# *APOE4* impact on soluble and insoluble tau pathology is mostly influenced by amyloid-beta

**DOI:** 10.1101/2024.09.20.24314064

**Authors:** Claudia Cicognola, Gemma Salvadó, Ruben Smith, Sebastian Palmqvist, Erik Stomrud, Tobey Betthauser, Sterling Johnson, Shorena Janelidze, Niklas Mattsson-Carlgren, Oskar Hansson, Alexa Pichet Binette

## Abstract

The *APOE4* allele is the strongest genetic risk factor for sporadic Alzheimer’s disease (AD). While *APOE4* is strongly associated with amyloid-beta (Aβ) accumulation, its relationship with tau accumulation is less understood. Studies evaluating the role of *APOE4* on tau accumulation have shown conflicting results, particularly regarding the independence of these associations from Aβ load. To clarify the relations between *APOE4,* Aβ and tau, we examined three independent longitudinal cohorts (the Swedish BioFINDER-1, BioFINDER-2 and WRAP cohorts) in which participants had cross-sectional and longitudinal measures of tau tangles (tau-PET; temporal meta-ROI and entorhinal) or soluble p-tau (p-tau217), Aβ-PET and *APOE* genotype. The study included a total of 1370 cognitively unimpaired (CU) and 449 mild cognitive impairment (MCI) subjects, followed longitudinally with tau-PET and p-tau217. *APOE4* carriers accounted for 40.2-50% of the cohorts. Different linear regressions (cross-sectional) and linear mixed-effect models (longitudinal) with tau measures as outcomes were fitted to test the effect of *APOE4* as independent predictor, as well as in combination with baseline Aβ load or the interaction term between *APOE4* and Aβ load. All models included age, sex and cognitive status as covariates.

We found no independent effects of the *APOE4* carriership on insoluble tau aggregates in either cohort (BioFINDER-2 or WRAP), both on cross-sectional and longitudinal tau-PET levels in the temporal meta-ROI, when Aβ was present in the model (p=0.531-0.949). Aβ alone was the best predictor of insoluble tau accumulation, and there was no interaction between *APOE4* and Aβ on tau-PET. Similarly, no independent effects of the *APOE4* carriership on baseline (p=0.683-0.708) and longitudinal (p=0.188-0.570) soluble p-tau217 were observed when Aβ was included in the model in BioFINDER-1 and WRAP. No interaction between *APOE4* and Aβ on soluble p-tau217 was observed. Furthermore, mediation analysis revealed that Aβ load fully mediated most associations between *APOE4* and tau (46-112%, either cross-sectional or longitudinal tau-PET or soluble p-tau217). In the largest cohort (BioFINDER-2), looking at *APOE4* groups based on the number of ε4 alleles, we found an interaction between APOE4 homozygotes only and Aβ on tau-PET levels at baseline and over time.

In conclusion, although *APOE4* is strongly associated with Aβ aggregation, it seems to be minimally associated with longitudinal changes in soluble or insoluble p-tau levels at a given level of Aβ pathology, confirming the primacy of Aβ in driving tau pathology.

## Introduction

The *APOE4* allele is the strongest genetic risk factor for sporadic Alzheimer’s disease (AD) [1–4]. Associations between *APOE4* genotype and amyloid β (Aβ) are also well-established, with carriers being more likely to be Aβ-positive and accumulating Aβ earlier in life than non-carriers [5–8]. Following the deposition of Aβ plaques in the brain, the deposition of neocortical tau aggregates ensues years later [9]. However, associations between *APOE* and longitudinal accumulation of soluble and insoluble hyperphosphorylated tau (p-tau) remain unclear. Some studies suggest that the presence of the ε4 allele potentiates the effects of Aβ on accumulation of insoluble tau aggregates and tau accumulation might be accelerated in ε4 carriers at lower Aβ levels [10–15]. However, others found no additional effect of *APOE4* on tau deposition beyond the effects on Aβ [16, 17]. It is important to clarify the role of *APOE4* on the two core pathologies of AD, considering recent efforts to develop *APOE*-targeting therapies. By an improved understanding of which AD pathology is most influenced by *APOE4* allele, and at which levels, we can better define the optimal target populations and outcomes in trials evaluating *APOE*-modifying therapies [18, 19]. For example, if only Aβ accumulation, and not tau accumulation, is associated with *APOE* genotype, *APOE*-targeting therapies should preferably be evaluated in primary (or potentially secondary) prevention trials. However, if *APOE4* has a strong direct effect on tau accumulation as well, trials could also include later disease stages such as cognitively impaired AD populations.

In the present study, we therefore aimed to assess if the presence of the *APOE4* allele modified the associations between insoluble tau (as measured with tau-PET) or soluble p-tau217 (as measured in CSF or plasma) and Aβ pathology, and whether *APOE4* had Aβ-independent effects on cross-sectional and longitudinal tau deposition. We examined cognitively unimpaired (CU) individuals and patients with mild cognitive impairment (MCI) from three independent longitudinal cohorts (BioFINDER-1, BioFINDER-2, WRAP). We put emphasis on the effects of *APOE4* on longitudinal tau measures, which allowed us to adjust for the fact that different participants are at different disease stages at baseline.

## Materials and methods

### Participants

The study participants of the first two cohorts belonged to the ongoing prospective Swedish BioFINDER study, namely BioFINDER-1 (NCT01208675) and BioFINDER-2 (NCT03174938), as previously described [20–22] (www.biofinder.se). All participants were recruited at the memory clinics of Skåne University Hospital and the Hospital of Ängelholm, Sweden. We included CU individuals and patients with MCI in the present study, for whom enrollment and follow-up visits were conducted between 2007 and 2015 for BioFINDER-1 and from 2017 to 2023 for BioFINDER-2. Briefly, participants included were 40 to 100 years old, had a Mini-Mental State Examination (MMSE) between 27 and 30 or 26 and 30 depending on age for cognitively unimpaired (CU) participants and between 24 and 30 for MCI, and were proficient in Swedish such that an interpreter was not necessary for cognitive testing. Exclusion criteria included severe somatic disease, current alcohol/substance misuse, refusing lumbar puncture or neuroimaging. Classification of cognitively unimpaired (CU) individuals followed the National Institute on Aging-Alzheimer’s Association criteria, i.e. not meeting criteria for Mild Cognitive Impairment (MCI) or dementia [23]. The CU group included cognitively healthy controls or individuals with subjective cognitive decline, who performed within normal ranges on a comprehensive cognitive test battery [24]. In BioFINDER-2, a group of younger controls (20-40 years old, n=41) was also included, which by design enrolled a high proportion of *APOE4* carriers (39%). In BioFINDER-2, MCI diagnosis was defined as performing below -1.5[standard deviation in any cognitive domain, employing a regression-based norm that accounted for age and education and z-scores based on Aβ-negative controls [25]. Cognitive domain z-scores were computed by averaging the z-scores of tests related to attention/executive function, verbal ability, memory, and visuospatial function. In BioFINDER-1, MCI determination was made by a senior neuropsychologist following comprehensive neuropsychological testing, as detailed elsewhere [26].

The Wisconsin Registry for Alzheimer’s Prevention (WRAP) cohort includes participants who were cognitively normal at enrollment, recruited from the population, enriched for parental history of AD and were between 40 and 65 years at baseline. Participants underwent neuropsychological evaluations on a biennial basis. More information about the recruitment of these participants can be found in [27]. For the present study, we refer to two different samples (“WRAP PET” and “WRAP plasma”) which were analyzed separately and share some participants, but not all as not everyone had both tau-PET and plasma measurements. For this analysis, baseline was considered as time of first tau-PET or plasma p-tau217 measurement, and by then a small proportion of participants had progressed to MCI. Cognitive status at each visit was determined by a consensus diagnosis and diagnosis of MCI was assigned based on National Institute on Aging-Alzheimer’s Association criteria without reference to biomarkers [28]. Complete inclusion and exclusion criteria are described elsewhere [29]. The WRAP PET sample included all participants who had completed Aβ- and tau-PET and T1-weighted MRI scans by April 2023. The WRAP plasma sample included participants who had an Aβ-PET and plasma p-tau217 levels, which was measured from visits that took place between August 2011 and January 2020.

All participants provided written informed consent prior to participation in the studies. The BioFINDER-studies received ethical approval from the Regional Ethical Committee in Lund, Sweden and the University of Wisconsin Institutional Review Board approved all WRAP study procedures. All research was completed in accordance with the Declaration of Helsinki.

### Image acquisition and processing

In the BioFINDER cohorts, PET images were were acquired as 4×5 min time frames on digital GE Discovery MI scanners. Aβ PET was done 90-110 minutes after the injection of ∼185 MBq [^18^F]flutemetamol and tau PET was done 70-90 minutes post injection of ∼370 MBq [^18^F]RO948. Briefly, images were motion corrected, averaged and co-registered to the closest T1 MPRAGE scan acquired on a Siemens MAGNETOM Prisma 3T MRI scanner at 1×1×1 mm isometric resolution. MRI scanning parameters have been described in detail previously. Cortical parcellation of the MRIs were performed using FreeSurfer, version 6.0. Standardized uptake value ratios (SUVRs) were then calculated using the cerebellum as reference region for Aβ and the inferior cerebellar cortex reference region for tau [30]. For the analysis, Aβ-PET measures were considered both as continuous SUVR and as binarized data using a cutoff derived from mixture modeling in the BioFINDER-2 cohort (1.03 SUVR) [31]. A neocortical meta-region of interest (ROI) for Aβ-PET (prefrontal, lateral temporal, parietal, anterior cingulate, and posterior cingulate/precuneus) was calculated [31]. For tau-PET, the regions of interest were (1) a temporal meta-ROI (bilateral entorhinal cortex, inferior and middle temporal cortices, fusiform gyrus, parahippocampal cortex, and amygdala) [30] and (2) the entorhinal cortex, given its previous associations with the *APOE4* allele [32–34].

In the WRAP PET cohort, all participants underwent Aβ- ([^11^C]-PiB; Pittsburgh compound B) and tau- ([^18^F]-MK-6240) PET imaging at the University of Wisconsin-Madison. Details regarding radiopharmaceutical production, acquisition protocols, as well as image reconstruction, processing and quantification of [^11^C]-PiB and [^18^F]-MK-6240 PET data have been described previously [35]. Aβ burden was assessed as a global average [^11^C]-PiB distribution volume ratio (DVR; Logan graphical analysis of the dynamic 0-70 minute acquisition using the cerebellum grey matter as the reference region), taken across eight bilateral cortical ROIs. A previously established threshold of DVR > 1.2 was used to define [^11^C]-PiB positivity [36] which corresponded to a centiloid of 21.6. For reporting, DVR values were linearly translated to centiloids [29]. Tau burden on the [^18^F]-MK-6240 tau-PET scan was quantified using SUVRs (70-90 min, inferior cerebellar grey matter reference region), in brain regions corresponding to the Harvard Oxford atlas. A temporal meta-ROI encompassing the amygdala, parahippocampal gyrus, inferior and middle temporal gyri was calculated, as well as the parahippocampal gyrus (corresponding to the entorhinal cortex in this atlas).

### CSF and plasma measurements

CSF samples in BioFINDER-1 were collected and handled according to a standardized protocol [37]. Samples were taken from non-fasting individuals by lumbar puncture at 3 different centers [38]. The CSF was centrifuged (2000*g*, +4°C, 10 minutes), aliquoted into 1 mL polypropylene tubes (Sarstedt AG & Co, Nümbrecht, Germany), and stored at −80°C. Samples went through 1 freeze-thaw cycle before the analysis when 200 μL was further aliquoted into LoBind tubes (Eppendorf Nordic A/S, Hørsholm, Denmark). Plasma samples from WRAP plasma were collected in EDTA tubes (BD 366643; Franklin Lakes, New Jersey, USA). Samples were mixed gently by inverting 10–12 times and were centrifuged 15 min at 2000*g* at room temperature within one hour of collection. Plasma samples were aliquoted into 2 mL cryovials (Wheaton Cryoelite W985863; Millville, New Jersey, USA). Aliquoted plasma was frozen at −80°C within 90 min and stored. CSF and plasma p-tau217 were measured using a Mesoscale Discovery immunoassay developed by Lilly Research Laboratories at Lund University. Samples were analyzed as previously described [39]. Briefly, samples were assayed in duplicates with biotinylated-IBA493 used as a capture antibody and SULFO-TAG-4G10-E2 as the detector. The assay was calibrated with a synthetic p-tau217 peptide.

### Statistics

Statistical analysis and data visualization were performed with R software version 4.2.3. P values <0.05 were considered significant. Participants were dichotomized in *APOE4* carriers and non-carriers based on whether they had at least one □4allele or no □4allele, respectively. In complementary analyses, we showed similar results based on the number of □4alleles (0 as non-carriers, 1 as *APOE4* heterozygotes, 2 as *APOE4* homozygotes) instead of the binary *APOE4* status, with a main focus on BioFINDER-2 given that its large sample size allowed to repeat all analyses. Separate linear regression models were used to determine the associations between baseline tau-PET and CSF/plasma p-tau217 measures with *APOE4*, Aβ and the interaction between *APOE4* and Aβ measures at baseline. Similarly, linear mixed-effect models were used to assess associations between longitudinal tau measures and the same parameters (*APOE4*, Aβ, and their interaction), using random slope and intercept. The “time” variable was calculated in years based on the dates when tau follow-up measures were performed. IDs that had missing data in one of the variables included in the lm and lme models were removed. Overall, we constructed four linear models for cross-sectional data and linear mixed-effect for longitudinal tau data as follows: 1) with only *APOE4* as predictor; 2) with only baseline Aβ PET as predictor; 3) with *APOE4* and baseline Aβ PET as predictors; 4) with the interaction *APOE4* x baseline Aβ PET as predictor. All continuous variables were normalized using the “scale()” function in R prior to this analysis. Aβ was used in continuous measures in all statistical models and was dichotomized only for visual representation. Lastly, mediation analysis was used to investigate the indirect effect of *APOE4* on tau variables through Aβ mediation. The analysis was conducted using the “mediation” package in R, using bootstrapping with 1000 resamples to obtain bias-corrected confidence intervals. All analyses were adjusted for age, sex, and cognitive status. For cross-sectional analyses, group differences were assessed with ANOVA, with Tukey’s Honest Significant Difference (HSD) post-hoc test for pairwise group comparisons.

### Data Availability

Pseudonymized BioFINDER data can be shared to qualified academic researchers after request for the purpose of replicating procedures and results presented in the study. In line with the EU General Data Protection Regulation (GDPR) legislation, a data transfer agreement must be established to share data. The agreement will include terms of how data is stored, protected, and accessed, and will define what the receiver can or cannot do. The agreement also must be approved by the Swedish Ethical Review Authority and Region Skåne. The data from the Wisconsin Registry for Alzheimer’s Prevention can be requested through online submission processes.

## Results

### Study cohorts and demographics

The study included a total of 1819 subjects, of which 1133 had longitudinal measurements. Samples came from three different cohorts, in which longitudinal measures of tau PET or soluble p-tau217 were available: the Swedish BioFINDER-2 study (baseline tau-PET, n=1048 with baseline tau PET; longitudinal tau-PET, n=629), the Swedish BioFINDER-1 study (baseline CSF p-tau217, n=237; longitudinal p-tau217, n=178), and WRAP, here subdivided in a “WRAP PET” cohort (baseline tau-PET, n=403; longitudinal tau-PET, n=199) and a “WRAP plasma” cohort (baseline plasma p-tau217, n=131; longitudinal p-tau217, n=127; Table 1, Supplementary table 1-2). In the BioFINDER studies, 31.9-40.9% of patients had MCI while the WRAP cohort was composed almost entirely of CU participants. *APOE4* carriers accounted for 40.2-50% of the cohorts. *APOE4* heterozygotes represented between 33 and 42% of the sample in the different cohorts, and *APOE4* homozygotes between 5 and 8% (Table 1). The detailed APOE genotypes across cohorts are listed in Supplementary Table 1. Across all cohorts, the Aβ-positive participants had elevated tau levels at baseline, but there was no difference in tau levels between the *APOE4* carriers or non-carriers within the Aβ-positive or negative groups (Supplementary Figure 1). The longitudinal subsets were followed up for an average time of 2.6-5.2 years, depending on the cohort. All demographic characteristics are detailed in Table 1, and in Supplementary Tables 2-3 where participants are broken down by *APOE4* status and cognitive status.

**Table 1.** Characteristics of the BioFINDER and WRAP cohorts.

| **Baseline** | **BioFINDER-2**<br>**(N=1048)** | **WRAP PET (N=403)** | **BioFINDER-1 (N=237)** | **WRAP plasma (N=131)** |
| --- | --- | --- | --- | --- |
| **Age, years** | 65.2 (13.2) | 68.1 (6.77) | 72.4 (5.41) | 62.5 (6.45) |
| **Sex, n (% female)** | 528 (50.4%) | 275 (68.2%) | 118 (49.8%) | 88 (67.2%) |
| **Education, years** | 12.9 (3.63) | 16.2 (2.67) | 11.6 (3.34) | 16.4 (2.43) |
| **Diagnosis, CU/MCI (% MCI)** | 714/334 (31.9%) | 388/15 (3.7%) | 140/97 (40.9%) | 128/3 (2.3%) |
| **APOE ε4 carriers, n (%)** | 524 (50.0%) | 162 (40.2%) | 101 (42.6%) | 59 (45.0%) |
| **Number of APOE ε4 alleles**<br>**0/1/2 (%)** | 524/441/83<br>(50/42/8%) | 241/132/30<br>(60/33/7%) | 136/83/18<br>(57/35/8%) | 72/52/7<br>(55/40/5%) |
| **Tau-PET temporal meta-ROI**<br>**SUVR<sup>1</sup>** | 1.24 (0.30) | 1.1 (0.23) | - | - |
| **Global Aβ-PET<sup>2</sup>** | 1.11 (0.30) | 18.98 (32.91) | 1.22 (0.33) | 25.3 (36.1) |
| **% Aβ+** | 361 (34.4%) | 23.1% | 54.4% | 31.3% |
| **P-tau217, pg/mL<sup>3</sup>** | - | - | 17.3 (22.9) | 0.3 (0.14) |
| **Longitudinal** | **BioFINDER-2 (N=629)** | **WRAP PET (N=199)** | **BioFINDER-1 (N=178)** | **WRAP plasma (N=127)** |
| **Length follow-up, years** | 2.6 (0.99) | 2.8 (1.01) | 3.7 (1.27) | 5.2 (1.72) |

Data are presented as mean (standard deviation) unless specified otherwise.<sup>1</sup>In BioFINDER-2 the tracer is RO948 and in WRAP it is MK6240;
<sup>2</sup>In the BioFINDER studies, Aβ-PET is in SUVR, in WRAP it is Centiloids; <sup>3</sup>In BioFINDER-1 it is CSF p-tau217 and in WRAP it is plasma p-tau217. Abbreviations: Aβ = beta-amyloid; APOE ε4= apolipoprotein E genotype; CSF= cerebrospinal fluid; CU= cognitively unimpaired; MCI= mild cognitive impairment; PET= positron emission tomography; SUVR= standardized uptake value ratio.

### *APOE4* effects on deposition and accumulation of insoluble tau are explained by the presence of Aβ

We first evaluated whether *APOE4* carriership was associated with the deposition (cross-sectional) and accumulation (longitudinal) of insoluble tau measured with tau-PET in BioFINDER-2 and WRAP PET, and, if present, whether such associations were influenced by the levels of Aβ pathology.

Looking at cross-sectional tau-PET in the temporal meta-ROI, in both cohorts, *APOE4* carriership was only associated with higher tau-PET burden in models that did not include other main predictors. In all models that included Aβ-PET load, Aβ was always associated with tau-PET levels, the effect of *APOE4* was no longer significant and there were no interactions between Aβ and *APOE4* (Table 2, Figure 1a-c). Further, in both cohorts the model explaining the highest variance was the one including only Aβ-PET (R^2^=0.364 in BioFINDER-2 and 0.333 in WRAP) and there were no improvements of the models by adding *APOE4* as covariate or the interaction between Aβ and *APOE4* (Table 2). Results focusing on tau-PET in the entorhinal cortex were different: in BioFINDER-2, there was a significant interaction between *APOE4* carriership and Aβ (b=0.166, p<0.001), such that for a given level of Aβ, *APOE4* carriers had higher tau-PET burden in the entorhinal cortex than non-carriers (Figure 1b and Supplementary Table 4). However, such an interaction was not present in WRAP PET, where results were similar as those in the temporal meta-ROI (Supplementary Table 4).

**Figure 1.**
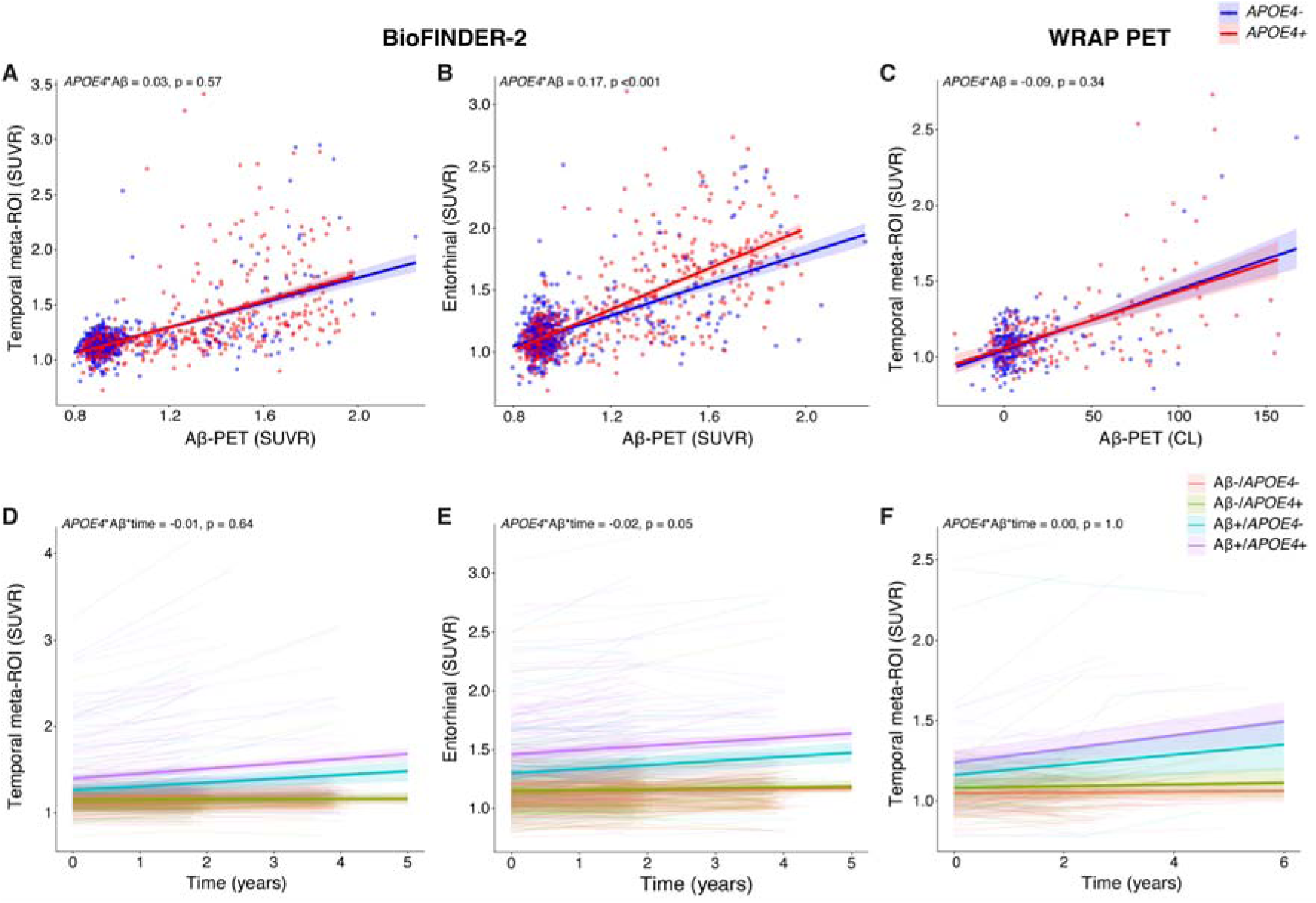
APOE4 and Aβ interaction on cross-sectional and longitudinal insoluble tau (tau PET SUVR) in BioFINDER-2 and WRAP PET. **a-c)** Scatter plot for visualization of the relationship between tau-PET SUVR and Aβ-PET SUVR at baseline divided by APOE □4status in BioFINDER-2 **(a-b)** and WRAP PET **(c). d-f)** Longitudinal tau-PET SUVR, groups divided by baseline Aβ PET status combined with APOE4 genotype in BioFINDER-2 **(c-d)** and WRAP PET **(d).** For visualization purposes, we created 4 groups based on Aβ status and APOE □4status, but continuous Aβ-PET SUVR was used in the statistical models. All statistical results are reported in Table 2.

**Table 2.** Effects of APOE4 and Aβ on cross-sectional and longitudinal insoluble tau (tau-PET SUVR)

| Model | Variable(s) | Baseline |  |  |  |  |  | Longitudinal |  |  |  |  |  |
| --- | --- | --- | --- | --- | --- | --- | --- | --- | --- | --- | --- | --- | --- |
|  |  | BioFINDER-2 (N=1048) |  |  | WRAP PET (N=403) |  |  | BioFINDER-2 (N=629) |  |  | WRAP PET (N=199) |  |  |
|  |  | b(SE) | p | R<sup>2</sup> | b(SE) | p | R<sup>2</sup> | b(SE) | p | AIC | b(SE) | p | AIC |
| 1 | APOE4 | 0.308<br>(0.057) | <0.001 | 0.162 | 0.368<br>(0.096) | <0.001 | 0.139 | 0.066<br>(0.014) | <0.001 | 1592.3 | 0.038<br>(0.025) | 0.127 | 906.1 |
| 2 | Aβ-PET | 0.552<br>(0.029) | <0.001 | 0.364 | 0.508<br>(0.044) | <0.001 | 0.333 | 0.093<br>(0.006) | <0.001 | 1411.9 | 0.067<br>(0.011) | <0.001 | 818.4 |
| 3 | APOE4 | 0.003<br>(0.052) | 0.949 | 0.364 | 0.025<br>(0.091) | 0.785 | 0.332 | 0.008<br>(0.013) | 0.531 | 1426.0 | 0.008<br>(0.025) | 0.737 | 829.2 |
| 3 | Aβ-PET | 0.551<br>(0.030) | <0.001 | | 0.503<br>(0.047) | <0.001 | | 0.092<br>(0.006) | <0.001 | | 0.065<br>(0.012) | <0.001 | |
| 4 | APOE4 | 0.006<br>(0.052) | 0.910 | 0.363 | 0.020<br>(0.091) | 0.829 | 0.332 | 0.006<br>(0.013) | 0.625 | 1429.5 | 0.009<br>(0.025) | 0.729 | 840.4 |
| 4 | Aβ-PET | 0.531<br>(0.046) | <0.001 | | 0.562<br>(0.077) | <0.001 | | 0.097<br>(0.012) | <0.001 | | 0.064<br>(0.016) | <0.001 | |
| 4 | APOE4 x<br>Aβ-PET | 0.031<br>(0.054) | 0.567 | | -0.091<br>(0.095) | 0.336 | | -0.006<br>(0.014) | 0.642 | | 0.000<br>(0.023) | 0.996 | |

Results from linear regression (baseline) and linear mixed-effects (longitudinal) models in BioFINDER-2 and WRAP PET with tau-PET SUVR as outcome. Models were constructed as follows: 1) with only APOE as predictor; 2) with only baseline Aβ-PET as predictor; 3) with APOE4 and baseline Aβ-PET as predictors; 4) with APOE4, baseline Aβ-PET and the interaction APOE4 x baseline Aβ-PET as predictor. All models were corrected for age, sex and cognitive status. For the linear mixed-effect models, the standardized beta reported correspond to the variable\*time. Aβ = beta-amyloid, B(SE)= standardized estimate with standard error; p= p-value; R<sup>2</sup> = R-squared, AIC= Akaike information criterion.

When using longitudinal tau-PET in the temporal meta-ROI as the outcome (BioFINDER-2 and WRAP PET), *APOE4* carriership was significantly associated with faster accumulation insoluble tau aggregates only in BioFINDER-2, in model in which *APOE4* genotype was the main predictor (Table 2). Conversely, Aβ load at baseline was a significant predictor of insoluble tau accumulation in all models where it was included (b=0.064-0.097, p<0.001). The models including only Aβ as predictor had the best model fit (represented by the lowest Akaike information criterion, Table 2) in both cohorts. Similar to the cross-sectional results, there was no interaction effect of *APOE* x Aβ x time on insoluble tau accumulation in any of the cohorts (p=0.642 and 0.996, Table 2). We additionally compared the longitudinal trajectories of tau accumulation between 4 groups defined by baseline Aβ-PET status combined with *APOE4* genotype (Aβ-□4-, Aβ-□4+, Aβ+□4-, Aβ+□4+; Figure 1c-e). Using Aβ-□4-participants as the reference group, the only significant differences in trajectories were between the Aβ+ and Aβ-groups (p<0.001). There were no differences in trajectories between the Aβ-□4- and the Aβ-□4+ groups or between the Aβ+□4+ and the Aβ+□4-groups across the two cohorts (p=0.171-0.683; Figure 1c-e). Looking at associations with longitudinal tau-PET uptake in the entorhinal cortex showed similar results than the temporal meta-ROI, with a main effect of Aβ on longitudinal tau-PET, and no significant effect of *APOE4* in models including both Aβ and the genotype both in BioFINDER-2 and WRAP PET (Supplementary Table 4).

Mediation analyses showed that Aβ was fully mediating the relationship between *APOE4* and baseline tau-PET levels in the temporal meta-ROI (93-99% of the total effect) as well as between *APOE*4 and longitudinal change in tau-PET (48-95% of the total effect) in both cohorts (p=0.203-0.949 for direct effect of *APOE4*, Supplementary Figure 2a-d). On cross-sectional levels of tau-PET in entorhinal cortex, Aβ had a partial mediating effect in BioFINDER-2 (p=0.019 for the direct effect of APOE4), but a fully mediating effect on longitudinal levels (86%, p<0.001, Supplementary Figure 3).

### Soluble p-tau217 levels are differentially moderated by APOE4 at baseline and longitudinally

Next, we repeated similar analyses assessing if *APOE4* carriership affected the levels and accumulation of tau, focusing on soluble p-tau 217 in CSF (BioFINDER-1) or plasma (WRAP) as outcome.

In BioFINDER-1, *APOE4* was associated with higher baseline p-tau217 levels when it was the only main predictor (b=0.468, p<0.001, Table 3), but had no significant effect when Aβ-PET was present in the model. However, Aβ-PET was a significant predictor of p-tau217 in all models (b=0.612-0.679, p<0.001, Table 3), and there were no interactions between *APOE4* and Aβ-PET on p-tau217 levels (Figure 2a). In the WRAP plasma cohort, unlike all other cross-sectional results, *APOE4* was not associated with p-tau217 levels when it was the sole predictor, and there was a significant interaction between *APOE4* and Aβ-PET (b=-0.258, p=0.030), such that at a given level of Aβ-PET, *APOE4* non-carriers had higher p-tau217 levels (Table 3, Fig. 2b). However, this interaction was no longer significant when removing an extreme outlier (an *APOE4* non-carrier who had the highest Aβ- and tau-PET levels [b=-0.143, p=0.285]). Aβ-PET was associated to p-tau217 in all models where it was included (p<0.001, Table 3).

**Figure 2.**
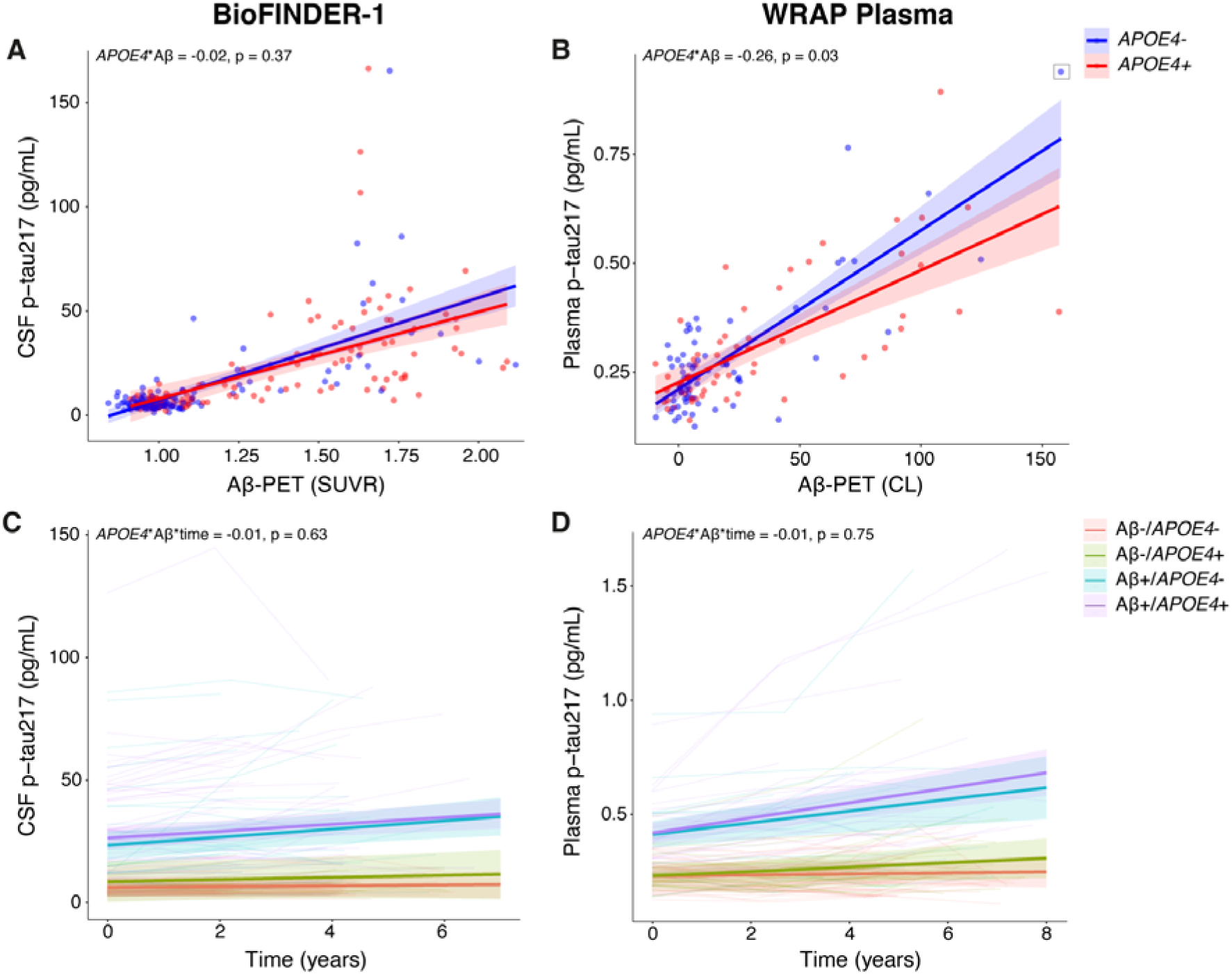
APOE4 and Aβ interaction on cross-sectional and longitudinal soluble p-tau217 in BioFINDER-1 and WRAP plasma. **a-b)** Scatter plot for visualization of the relationship between CSF **(a)** and plasma **(b)** p-tau217 and Aβ-PET SUVR at baseline divided by APOE4 status in BioFINDER-1 **(a)** and WRAP plasma **(b). c-d)** Longitudinal CSF **(c)** and plasma **(d)** p-tau217, groups divided by baseline Aβ-PET status combined with APOE4 genotype in BioFINDER-1 **(c)** and WRAP plasma **(d).** For visualization purposes, we created 4 groups based on Aβ status and APOE4 status, but continuous Aβ-PET SUVR was used in the statistical models. All statistical results are reported in Table 3.

**Table 3.** Effects of APOE4 and Aβ on cross-sectional and longitudinal soluble p-tau217.

| Model | Variable(s) | Baseline |  |  |  |  |  | Longitudinal |  |  |  |  |  |
| --- | --- | --- | --- | --- | --- | --- | --- | --- | --- | --- | --- | --- | --- |
|  |  | BioFINDER-1 (N=237) |  |  | WRAP plasma (N=131) |  |  | BioFINDER-1 (N=178) |  |  | WRAP plasma (N=127) |  |  |
|  |  | b(SE) | p | R<sup>2</sup> | b(SE) | p | R<sup>2</sup> | b(SE) | p | AIC | b(SE) | p | AIC |
| 1 | APOE4 | 0.468<br>(0.124) | **<0.001** | 0.138 | 0.324<br>(0.175) | 0.066 | 0.042 | 0.026<br>(0.018) | 0.134 | 657.6 | 0.058<br>(0.022) | **0.009** | 560.2 |
| 2 | Aβ-PET | 0.612<br>(0.056) | **<0.001** | 0.394 | 0.762<br>(0.059) | **<0.001** | 0.579 | 0.020<br>(0.009) | **0.024** | 569.2 | 0.048<br>(0.009) | **<0.001** | 458.8 |
| 3 | APOE4 | -0.048<br>(0.116) | 0.683 | 0.392 | -0.049<br>(0.120) | 0.686 | 0.576 | 0.011<br>(0.020) | 0.570 | 581.4 | 0.026<br>(0.020) | 0.194 | 470.3 |
| 3 | Aβ-PET | 0.623<br>(0.063) | **<0.001** | | 0.768<br>(0.061) | **<0.001** | | 0.017<br>(0.010) | 0.072 | | 0.045<br>(0.009) | **<0.001** | |
| 4 | APOE4 | -0.044<br>(0.116) | 0.708 | 0.391 | -0.046<br>(0.118) | 0.701 | 0.589 | 0.012<br>(0.020) | 0.548 | 593.7 | 0.026<br>(0.020) | 0.188 | 483.6 |
| 4 | Aβ-PET | 0.679<br>(0.089) | **<0.001** | | 0.909<br>(0.878) | **<0.001** | | 0.022<br>(0.014) | 0.110 | | 0.048<br>(0.014) | **<0.001** | |
| 4 | APOE4 x<br>Aβ-PET | -0.105<br>(0.116) | 0.367 | | -0.258<br>(0.118) | **0.030** | | -0.009<br>(0.019) | 0.626 | | -0.006<br>(0.018) | 0.745 | |

Results from linear regression (baseline) and linear mixed-effects (longitudinal) models in BioFINDER-1 and WRAP plasma with soluble p-tau217 (CSF or plasma) as outcome. Models were constructed as follows: 1) with only APOE4 as predictor; 2) with only baseline Aβ-PET as predictor; 3) with APOE4 and baseline Aβ-PET as predictors; 4) with APOE4, baseline Aβ-PET and the interaction APOE4 x baseline Aβ-PET as predictor. All models were corrected for age, sex and cognitive status. For the linear mixed-effect models, the standardized beta reported correspond to the variable\*time. B(SE)= standardized estimate with standard error; p= p-value; R<sup>2</sup> = R-squared, AIC= Akaike information criterion.

When using longitudinal soluble p-tau217 levels as the outcome, the only significant association with longitudinal p-tau217 in BioFINDER-1 was with baseline Aβ-PET SUVR in the model where it was the main predictor (b=0.026, p=0.024, Table 3). When *APOE4* alone or both *APOE4* and Aβ-PET were present in the model, none of the longitudinal associations were significant, but the effect of Aβ-PET SUVR was at trend level (p=0.072, Table 3). Of note, if using instead dichotomous Aβ-PET status (+/-) as the predictor, Aβ-PET was associated with p-tau217 in all models where it was included (b=0.032-0.038, p<0.001-0.002, Supplementary Table 5). In WRAP, *APOE<u>4</u>* was associated with longitudinal p-tau217 only when it was the main predictor (b=0.058, p=0.009, Table 3). Aβ-PET was associated to p-tau217 in all models where it was included (p<0.001, Table 3). In both cohorts, there were no interactions between *APOE4* and Aβ-PET load on p-tau217 levels over time. The longitudinal trajectories of soluble p-tau217 in the 4 groups defined by dichotomous Aβ-PET status (+/-) combined with *APOE4* genotype showed no difference between the Aβ-□4- and the Aβ-□4+ group or between the Aβ+□4+ and Aβ+□4-group (p=0.585-0.673 in BioFINDER-1; p=0.174-0.377 in WRAP, Figure 2c-d).

Mediation analyses showed that baseline Aβ-PET fully mediated the association between *APOE4* and cross-sectional p-tau217 levels in both cohorts and with longitudinal p-tau217 in the BioFINDER-1 cohort (110-112% of the total effect; Supplementary Figure 4a-c). However, *APOE4* had a significant direct effect on longitudinal p-tau217 in WRAP (b=0.008, p=0.034), and Aβ-PET had a partial mediating effect (b=0.007, p=0.006, 46% of mediation Supplementary Figure 4d).

### Differential associations based on number of *APOE4* alleles in BioFINDER-2

Lastly, we investigated similar associations based on the number of *APOE4* alleles (0, 1 or 2) instead of the dichotomous *APOE4* carriership. Such analyses were focussed on BioFINDER-2 given the largest sample size, which allowed to have 83 *APOE4* homozygotes. Looking at cross-sectional association with tau-PET, results were different between the temporal meta-ROI and the entorhinal cortex. In the former, there was a significant interaction between Aβ-PET and the *APOE4* homozygotes compared to non-carriers or *APOE4* heterozygotes (Fig. 3a, Supplementary Table 6). The interaction between Aβ-PET and the *APOE4* heterozygotes and non-carriers was not significant (Supplementary Table 6). Overall, for a given level of Aβ, only the *APOE4* homozygotes had higher tau burden compared to the two other groups. In the entorhinal cortex, the interaction between Aβ-PET and the *APOE4* was significant for all *APOE4* comparisons (Fig. 3b, Supplementary Table 6). Looking at associations with longitudinal tau-PET in the same regions, in the temporal meta-ROI, results were consistent with the interactions seen at the cross-sectional level: the *APOE4* homozygotes accumulated faster tau compared to the two other groups for a given level of Aβ (*APOE4* homozygotes*Aβ*time compared to non-carriers or heterozygotes with p=0.036 and p=0.002, Figure 3c, Supplementary Table 6). In the entorhinal cortex, there was not any significant associations with the genotypes, neither *APOE4* groups*time or *APOE4* groups*Aβ*time across the different models. The main effect on longitudinal tau uptake in the entorhinal cortex was higher levels of Aβ at baseline (Aβ*time: p<0.001, Figure 3d, Supplementary Table 6). All other cohorts had a smaller sample size and thus were not investigated in detail beyond APOE4 dichotomization. Still, in all cross-sectional measures, none of the other cohorts showed significant interactions between the three *APOE4* groups (Supplementary Figure 5).

**Figure 3.**
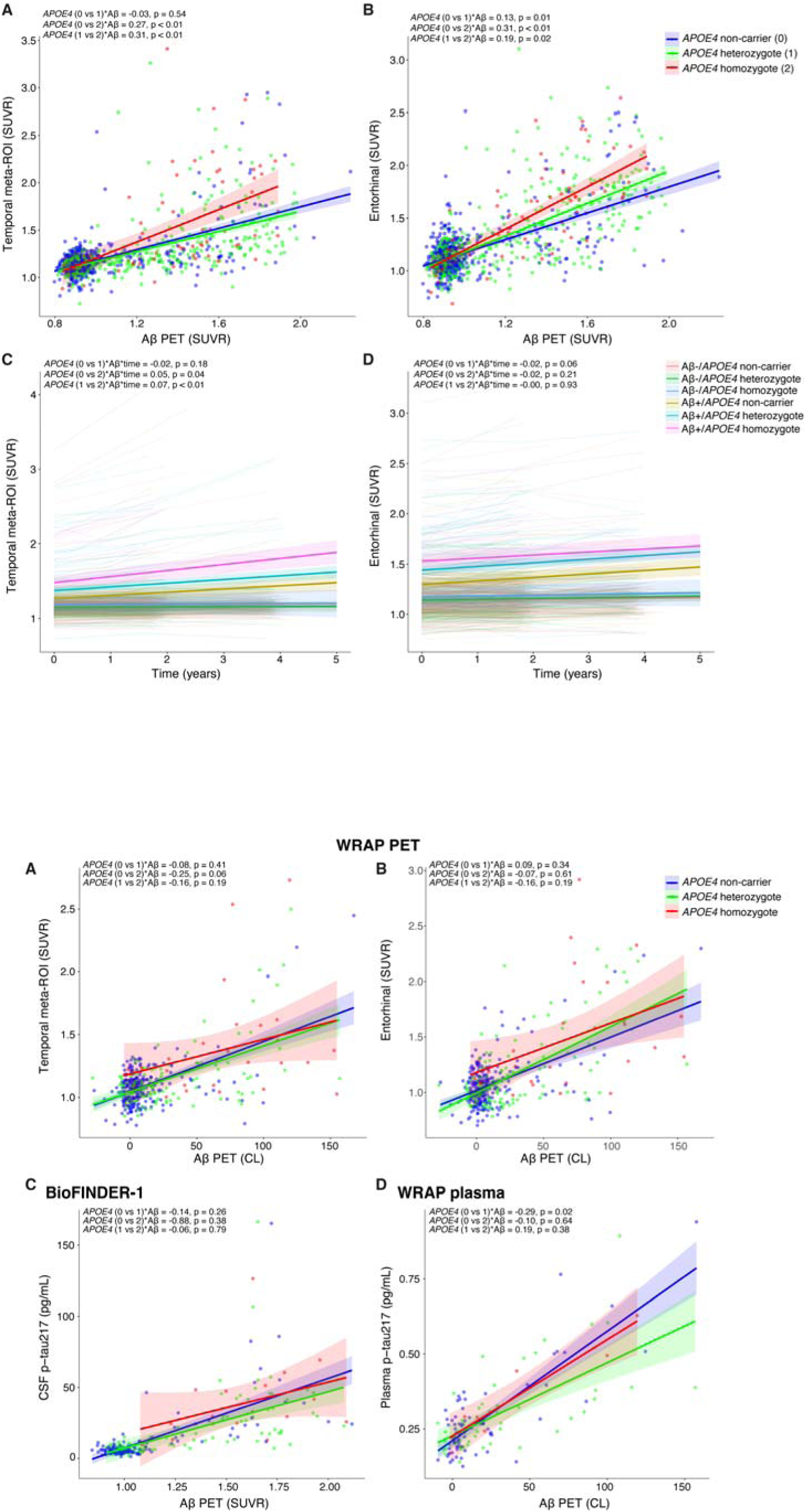
Interaction between APOE4 groups based on the number of ε4 alleles and Aβ on cross-sectional and longitudinal insoluble tau-PET in BioFINDER-2. **a-b)** Scatter plot for visualization of the relationship between tau-PET SUVR (temporal meta-ROI in a and entorhinal cortex in **b)** and Aβ-PET SUVR split between three APOE4 groups. 0 corresponds to non-carriers, 1 corresponds to APOE4 heterozygotes; 2 corresponds to APOE4 homozygotes **c-d)** Longitudinal tau-PET SUVR, groups divided by baseline Aβ PET status combined with the three different APOE4 groups. For visualization purposes, we created 6 groups based on Aβ status and number of □4 alleles, but continuous Aβ-PET SUVR was used in the statistical models. All statistical results are reported in Supplementary Table 5.

## Discussion

Our study aimed to investigate the interplay of *APOE4* genotype and Aβ on both baseline tau burden and longitudinal accumulation in the context of the AD continuum. Our results, which were very consistent in three cohorts across cross-sectional and longitudinal tau-PET or soluble p-tau measures, challenge the hypothesis that *APOE4* significantly influences tau accumulation in the temporal meta-ROI independently of Aβ load. In cross-sectional analyses, in the largest cohort we observed a significant interaction between Aβ and *APOE4* carriership on tau-PET uptake in the medial temporal lobe. In longitudinal analyses, *APOE4* carriership was not a significant predictor of accumulation of insoluble tau aggregates or soluble p-tau217 when Aβ load was also included as a predictor in the models across all cohorts. The tau accumulation trajectories of the Aβ+/ε4+ for group was not significantly steeper than the ones of the Aβ+/ε4-group. No major, direct role of *APOE4* on tau was further supported by the mediation analyses, which showed that in almost all cases, Aβ fully mediated the effects of *APOE* on tau deposition and accumulation. Still, we should mention that when looking at groups split based on the number of *APOE4* alleles in the largest cohort, the *APOE4* homozygotes showed a significant interaction with Aβ on tau-PET in the temporal meta-ROI, both at the cross-sectional and longitudinal level, which might point to different associations in this population particularly at risk of AD. Taken together, these findings suggest that the main way *APOE4* carriership might affect tau accumulation is through Aβ load, both at baseline and over time.

The current findings converge with those of previous studies where the effect of the ε4 allele on the baseline load or the accumulation (i.e., change in tau-PET) of insoluble tau in the neocortical temporal regions were entirely mediated by Aβ load [16, 40]. Likewise, another study has shown that ε4 carriers did not have higher rates of accumulation of insoluble tau aggregates [17, 41]. A recent study on plasma NTA tau (a N-terminal soluble tau fragment associated to AD) showed that ε4 carriers do not have higher plasma tau concentrations compared to non-carriers [42]. However, results differ slightly when looking at insoluble tau in the medial temporal lobe. In BioFINDER-2, the largest cohort included in this paper, we found an interaction such that for a given level of Aβ, *APOE* carriers had higher tau levels than non-carriers. Such a result is also in line with previous studies done across the AD continuum [11, 32, 33]. We did not see this interaction in the WRAP cohort, probably due to the lower tau levels in that cohort, composed of mostly cognitively normal older adults. Still, we are cautious not to infer Aβ-independent pathway through which *APOE4* affects insoluble tau accumulation [10, 15], given that we did not see an Aβ-independent over time, nor when looking beyond *APOE4* dichotomization. The two previous studies suggesting an *APOE4* effect independent of Aβ have several differences from ours, including smaller sample sizes and the use of different statistical methods (dichotomization of tau and Aβ measures, logistic regression). Dichotomization of Aβ, specifically, might mask the fact that the *APOE4* carriers might have higher amounts of Aβ pathology since the aggregation of Aβ starts at an earlier age, which is an important consideration in interpreting the literature.

In BioFINDER-2, when investigating three groups based on the number of *APOE4* alleles, the different associations seen in the temporal meta-ROI compared to the medial temporal lobe (an early region of tau deposition) might give credence to the recent idea that *APOE4* homozygotes are a “distinct genetic form of AD” [5]. In the temporal lobe composite region, both at baseline and over time, the *APOE4* homozygotes diverged both from heterozygotes and non-carriers, while the two latter groups behaved similarly. For the same level of Aβ, only the homozygotes showed higher tau aggregates or faster accumulation of aggregates, potentially hinting at more advanced pathology specifically in this genetic form. On the other hand, in the medial temporal lobe, at baseline, we observed a more “dose-dependent” effect of the number of *APOE4* interacting with Aβ. However, over time, the larger effect on accumulation of tau aggregates in this early tau region was higher levels of Aβ at baseline, and not *APOE* genotype. We postulate that the “dose-dependent” effect on baseline levels might rather represent individuals being at different stages of the disease.

The current results have important implication for therapeutics targeting *APOE* [19], in such that these therapies might likely affect the rate of Aβ aggregation, but not directly tau aggregation. We thus speculate that *APOE* therapies might work better if initiated during preclinical AD stages to stop or reduce the accumulation of Aβ, before Aβ has become widespread and drives tau accumulation in the neocortex. Supporting this notion, a recent study based on the ADNI cohort found that *APOE4* was an important factor in the progression from A-T- to A+T-, but only had a marginal effect in the progression from A+T- to A+T+ [43]. Consistent with these findings on tau, a study that examined effects of *APOE4* on cognition also did not find any effects of *APOE4* on cognition after accounting for Aβ load [44].

Here we provided a comprehensive assessment of tau dynamics based on a large sample size, longitudinal data, three independent cohorts and overall consistent results across different tau-PET tracers and soluble p-tau217 measures (both CSF and plasma). We acknowledge that the cohorts studied here were on average younger and were enriched by design to include a higher proportion of *APOE4* carriers, which might explain part of the differences with previous literature. The current results based on the three *APOE4* groups are also derived from only one cohort, and it will be interesting to see if similar associations are present in other large samples. The observational nature of the study does not allow to establish direct causation of our findings. Almost all participants included in the current study were White, and the Aβ-dependent or independent effect of *APOE4* on tau will be important to study in ethnically diverse populations, particularly given the different impact of *APOE* on AD pathology in different ethnicities [45].

In summary, our study sheds light on the intricate relationships between *APOE4*, Aβ, and tau in AD. In showing that overall Aβ load surpasses the impact of *APOE4* on tau accumulation, our findings strengthen the links between *APOE4* and Aβ and further investigating the genetic and molecular pathways underlying such links might help to find therapeutics to postpone the disease onset. We found that most effect of *APOE4* on tau accumulation in humans is mediated by Aβ load, with potential differences in *APOE4* homozygotes, which will need to be investigated further in other large samples.

