## Supplementary material for "*APOE4* impact on soluble and insoluble tau pathology is mostly influenced by amyloid-beta"

**Supplementary Table 1. *APOE* genotypes by cohort**
**Supplementary Table 2. Characteristics of the BioFINDER-2 and WRAP PET cohorts split by *APOE4* status and cognitive status**

**Supplementary Table 3. Characteristics of the BioFINDER-1 and WRAP plasma cohorts split by *APOE4* status and diagnosis**

**Supplementary Table 4. Effects of *APOE4* and Aβ status entorhinal tau-PET uptake in BioFINDER-2 and WRAP PET**

**Supplementary Table 5. Effects of *APOE4* and Aβ status on CSF p-tau217 in BioFINDER-1**

**Supplementary Table 6. Effects of *APOE4* based on the number of ε4 alleles and Aβ status tau-PET uptake in BioFINDER-2**

**Supplementary Figure 1. Cross-sectional tau levels based on Aβ and *APOE4* groups**

**Supplementary Figure 2. Mediation of Aβ on *APOE4* and temporal meta-ROI tau PET**

**Supplementary Figure 3. Mediation of Aβ on *APOE4* and entorhinal tau PET**

**Supplementary Figure 4. Mediation of Aβ on *APOE4* and soluble p-tau217**

**Supplementary Figure 5. Interaction between *APOE4* based on the number of ε4 alleles and Aβ on tau-PET and p-tau217**

**Supplementary Table 1. *APOE* genotypes by cohort**

|  | **BioFINDER-2** | **WRAP PET** | **BioFINDER-1** | **WRAP plasma** |
| --- | --- | --- | --- | --- |
| ***E2/E2*** | 4 | - | 1 | - |
| ***E2/E3*** | 79 | 38 | 21 | 10 |
| ***E2/E4*** | 31 | 12 | 7 | 5 |
| ***E3/E3*** | 441 | 203 | 114 | 62 |
| ***E3/E4*** | 410 | 120 | 76 | 47 |
| ***E4/E4*** | 83 | 30 | 18 | 7 |

Number of participants for each combination of *APOE* genotype

**Supplementary Table 2. Characteristics of the BioFINDER-2 and WRAP PET cohorts split by *APOE4* status and cognitive status**

|  | **BioFINDER-2** | | | | **WRAP PET** | | | |
| --- | --- | --- | --- | --- | --- | --- | --- | --- |
|  | ***APOE4-*** | | ***APOE4+*** | | ***APOE4-*** | | ***APOE4+*** | |
|  | **CU** | **MCI** | **CU** | **MCI** | **CU** | **MCI** | **CU** | **MCI** |
| **N** | 371 | 153 | 343 | 181 | 237 | 4 | 151 | 11 |
| **Age, years** | 63.2 (14.8) | 71.8 (8.99) | 61.2 (13.20) | 71.5 (7.22) | 68.9 (6.93) | 71.0 (4.52) | 66.6 (6.34) | 71.8 (5.83) |
| **Sex, % female** | 53.4% | 38.6% | 54.5% | 46.4% | 70.0% | 0% | 66.9% | 72.7% |
| **Education, years** | 12.9 (3.11) | 12.9 (4.44) | 13.1 (3.63) | 12.5 (3.84) | 16.3 (2.80) | 15.0 (3.46) | 16.0 (2.45) | 16.4 (2.42) |
| **% Aβ+** | 11.9% | 38.6% | 31.2% | 83.4% | 11.0% | 50.0% | 36.4% | 90.9% |
| **Tau-PET SUVR^1^** | 1.2 (0.16) | 1.3 (0.34) | 1.19 (0.21) | 1.48 (0.44) | 1.1 (0.18) | 1.1 (0.08) | 1.2 (0.21) | 1.6 (0.63) |

Data are presented as mean (standard deviation) unless specified otherwise. *APOE4*- are non-carriers and *APOE4*+ have at least one ε4 allele. ^1^In BioFINDER-2 the tracer is RO948 and in WRAP it is MK6240.

Abbreviations: Aβ= beta-amyloid; *APOE4*= apolipoprotein E genotype; CU= cognitively unimpaired; MCI= mild cognitive impairment; PET= positron emission tomography; SUVR= standardized uptake value ratio.

**Supplementary Table 3. Characteristics of the BioFINDER-1 and WRAP plasma cohorts split by *APOE4* status and diagnosis**

|  | **BioFINDER-1** | | | | **WRAP plasma** | | |
| --- | --- | --- | --- | --- | --- | --- | --- |
|  | ***APOE4-*** | | ***APOE4+*** | | ***APOE4-*** | | ***APOE4+*** |
|  | **CU** | **MCI** | **CU** | **MCI** | **CU** | **MCI** | **CU** |
| **N** | 87 | 49 | 55 | 48 | 69 | 3 | 59 |
| **Age, years** | 73.6 (5.22) | 71.4 (5.57) | 72.0 (5.69) | 71.7 (5.05) | 62.9 (6.26) | 70.1 (3.92) | 61.6 (6.55) |
| **Sex, % female** | 60.9% | 30.6% | 56.6% | 41.7% | 71.0% | 0% | 66.1% |
| **Education, years** | 11.9 (3.48) | 11.4 (3.45) | 11.7 (3.01) | 11.0 (3.33) | 16.5 (2.65) | 15.3 (3.06) | 16.3 (2.14) |
| **% Aβ+** | 29.9% | 46.9% | 64.2% | 95.8% | 20.3% | 33.3% | 44.1% |
| **P-tau217, pg/mL**^1^ | 9.2 (11.2) | 16.8 (27.8) | 16.4 (19.9) | 33.4 (28.0) | 0.3 (0.13) | 0.4 (0.31) | 0.3 (0.15) |

Data are presented as mean (standard deviation) unless specified otherwise. *APOE4*- are non-carriers and *APOE4*+ have at least one ε4 allele. ^1^In BioFINDER-1 it is CSF p-tau217 and in WRAP it is plasma p-tau217.

Abbreviations: Aβ= beta-amyloid; *APOE4*= apolipoprotein E genotype; CU= cognitively unimpaired; MCI= mild cognitive impairment; PET= positron emission tomography.

**Supplementary Table 4. Effects of *APOE4* and Aβ status entorhinal tau-PET uptake in BioFINDER-2 and WRAP PET**

|  | | **Baseline** | | | | | | **Longitudinal** | | | | | |
| --- | --- | --- | --- | --- | --- | --- | --- | --- | --- | --- | --- | --- | --- |
|  |  | **BioFINDER-2 (N=1048)** | | | **WRAP PET (N=403)** | | | **BioFINDER-2 (N=629)** | | | **WRAP PET (N=199)** | | |
| Model | **Variable(s)** | b(SE) | p | R^2^ | b(SE) | p | R^2^ | b(SE) | p | AIC | b(SE) | p | AIC |
| 1 | *APOE*4 | 0.450 (0.054) | **<0.001** | 0.250 | 0.470 (0.096) | **<0.001** | 0.152 | 0.036 (0.009) | **<0.001** | 1901.8 | 0.032 (0.025) | 0.204 | 883.2 |
| 2 | Aβ-PET | 0.638 (0.025) | **<0.001** | 0.503 | 0.567 (0.042) | **<0.001** | 0.382 | 0.043 (0.005) | **<0.001** | 1687.3 | 0.042 (0.011) | **<0.001** | 814.6 |
| 3 | *APOE*4 | 0.108 (0.046) | **0.019** | 0.505 | 0.095 (0.087) | 0.278 | 0.382 | 0.009 (0.009) | 0.312 | 1699.0 | 0.012 (0.025) | 0.630 | 823.5 |
| 3 | Aβ-PET | 0.618 (0.027) | **<0.001** |  | 0.549 (0.045) | **<0.001** |  | 0.041 (0.005) | **<0.001** |  | 0.041 (0.012) | **<0.001** |  |
| 4 | *APOE*4 | 0.122 (0.046) | **0.008** | 0.510 | 0.100 (0.087) | 0.256 | 0.382 | 0.006 (0.009) | 0.546 | 1698.3 | 0.013 (0.025) | 0.60433 | 835.3 |
| 4 | Aβ-PET | 0.511 (0.040) | **<0.001** |  | 0.492 (0.074) | **<0.001** |  | 0.057 (0.009) | **<0.001** |  | 0.045 (0.016) | **0.005** |  |
| 4 | *APOE4* x Aβ-PET | 0.166 (0.047) | **<0.001** |  | 0.088 (0.091) | 0.333 |  | -0.021 (0.011) | 0.050 |  | -0.009 (0.023) | 0.706 |  |

Results from linear regression (baseline) and linear mixed-effects (longitudinal) models in BioFINDER-2 and WRAP PET with tau-PET in the entorhinal as outcome. Models were constructed as follows: 1) with only *APOE4* as predictor; 2) with only baseline Aβ-PET as predictor; 3) with *APOE4* and baseline Aβ-PET as predictors; 4) with *APOE4*, baseline Aβ-PET and the interaction *APOE4* x baseline Aβ-PET as predictor. All models were corrected for age, sex and cognitive status. For the linear mixed-effect models, the standardized beta reported correspond to the variable*time. B(SE)= standardized estimate with standard error; p= p-value; R^2^ = R-squared, AIC= Akaike information criterion.

**Supplementary Table 5. Effects of *APOE4* and Aβ status on CSF p-tau217 in BioFINDER-1**

| **BioFINDER-1** | | **Baseline (N=237)** | | | **Longitudinal (N=178)** | | |
| --- | --- | --- | --- | --- | --- | --- | --- |
| Model | **Variable(s)** | b(SE) | p | R^2^ | b(SE) | p | AIC |
| 1 | *APOE*4 | 0.468 (0.124) | **<0.001** | 0.138 | 0.027 (0.018) | 0.134 | 657.6 |
| 2 | Aβ-PET status | 0.394 (0.060) | **<0.001** | 0.229 | 0.032 (0.008) | **<0.001** | 610.8 |
| 3 | *APOE*4 | 0.182 (0.128) | 0.157 | 0.232 | -0.003 (0.019) | 0.890 | 622.1 |
| 3 | Aβ-PET status | 0.355 (0.066) | **<0.001** |  | 0.033 (0.009) | **<0.001** |  |
| 4 | *APOE*4 | 0.164 (0.131) | 0.212 | 0.230 | -0.000 (0.019) | 0.989 | 633.7 |
| 4 | Aβ-PET status | 0.327 (0.080) | **<0.001** |  | 0.038 (0.012) | **0.002** |  |
| 4 | *APOE*4 x Aβ-PET status | 0.084 (0.136) | 0.538 |  | -0.013 (0.019) | 0.497 |  |

Results from linear regression (baseline) and linear mixed-effects (longitudinal) models in BioFINDER-1 for the effects of *APOE4* and binary Aβ-PET status on CSF p-tau217. Models were constructed as follows: 1) with only *APOE4* as predictor; 2) with only baseline Aβ-PET status as predictor; 3) with *APOE4* and baseline Aβ-PET status as predictors; 4) with *APOE4*, baseline Aβ-PET status and the interaction *APOE4* x baseline Aβ-PET status as predictor. All models were corrected for age, sex and diagnosis.
Abbreviations: Aβ= beta-amyloid; *APOE4*= apolipoprotein E genotype; b(SE)= standardized estimate with standard error; p= p-value; R^2^ = R-squared, AIC= Akaike information criterion.

**Supplementary Table 6. Effects of *APOE4* based on the number of ε4 alleles and Aβ status tau-PET uptake in BioFINDER-2**

| **BioFINDER-2** | | **Baseline (n=1048)** | | | | | | **Longitudinal (n=629)** | | | | | |
| --- | --- | --- | --- | --- | --- | --- | --- | --- | --- | --- | --- | --- | --- |
|  |  | **Temporal meta-ROI** | | | **Entorhinal cortex** | | | **Temporal meta-ROI** | | | **Entorhinal cortex** | | |
| Model | **Variable(s)** | b(SE) | p | R^2^ | b(SE) | p | R^2^ | b(SE) | p | AIC | b(SE) | p | AIC |
| 1 | *APOE*4 0 vs 1 allele | 0.218 (0.058) | **<0.001** | 0.184 | 0.378 (0.056) | **<0.001** | 0.264 | 0.051 (0.015) | **0.001** | 1594.5 | 0.035 (0.010) | **<0.001** | 1906.3 |
| 1 | *APOE*4 0 vs 2 alleles | 0.806 (0.108) | **<0.001** |  | 0.847 (0.102) | **<0.001** |  | 0.133 (0.026) | **<0.001** |  | 0.038 (0.018) | **0.032** |  |
| 1 | *APOE*4 1 vs 2 alleles | 0.588 (0.109) | **<0.001** |  | 0.469 (0.103) | **<0.001** |  | 0.082 (0.026) | **0.002** |  | 0.003 (0.018) | 0.864 |  |
| 2 | *APOE*4 0 vs 1 allele | -0.052 (0.053) | 0.331 | 0.374 | 0.072 (0.047) | 0.128 | 0.509 | -0.002 (0.013) | 0.893 | 1431.5 | 0.011 (0.010) | 0.268 | 1707.1 |
| 2 | *APOE*4 0 vs 2 alleles | 0.357 (0.100) | **<0.001** |  | 0.338 (0.087) | **<0.001** |  | 0.057 (0.023) | **0.014** |  | 0.001 (0.017) | 0.951 |  |
| 2 | *APOE*4 1 vs 2 alleles | 0.409 (0.100) | **<0.001** |  | 0.266 (0.085) | **0.002** |  | 0.059 (0.023) | **0.011** |  | -0.01 (0.017) | 0.579 |  |
| 2 | Aβ-PET | 0.538 (0.030) | **<0.001** |  | 0.610 (0.027) | **<0.001** |  | 0.090 (0.006) | **<0.001** |  | 0.042 (0.005) | **<0.001** |  |
| 3 | *APOE4* 0 vs 1 x Aβ-PET | -0.034 (0.055) | 0.536 | 0.380 | 0.127 (0.049) | **0.01** | 0.516 | -0.019 (0.014) | 0.181 | 1438.9 | -0.021 (0.011) | 0.062 | 1716.6 |
| 3 | *APOE4* 0 vs 2 x Aβ-PET | 0.274 (0.091) | **0.003** |  | 0.314 (0.081) | **<0.001** |  | 0.047 (0.022) | **0.036** |  | -0.022 (0.018) | 0.21 |  |
| 3 | *APOE4 1* vs 2 x Aβ-PET | 0.309 (0.087) | **<0.001** |  | 0.188 (0.077) | **0.015** |  | 0.066 (0.021) | **0.002** |  | -0.002 (0.016) | 0.926 |  |

Results from linear regression (baseline) and linear mixed-effects (longitudinal) models in BioFINDER-2 with tau-PET in the temporal meta-ROI and in the entorhinal as outcomes. The *APOE4* groups are coded based on the number of alleles: 0 corresponds to non-carriers, 1 corresponds to *APOE4* heterozygotes; 2 corresponds to *APOE4* homozygotes. Models were constructed as follows: 1) with only *APOE4* groups as predictor; 2) with *APOE4* groups and baseline Aβ-PET as predictors; 3) with the interaction of *APOE4* groups x baseline Aβ-PET as predictor. All models were corrected for age, sex and cognitive status. For the linear mixed-effect models, the standardized beta reported correspond to the variable*time. B(SE)= standardized estimate with standard error; p= p-value; R^2^ = R-squared, AIC= Akaike information criterion.

**Supplementary Figure 1. Cross-sectional tau levels based on Aβ and *APOE4* groups**

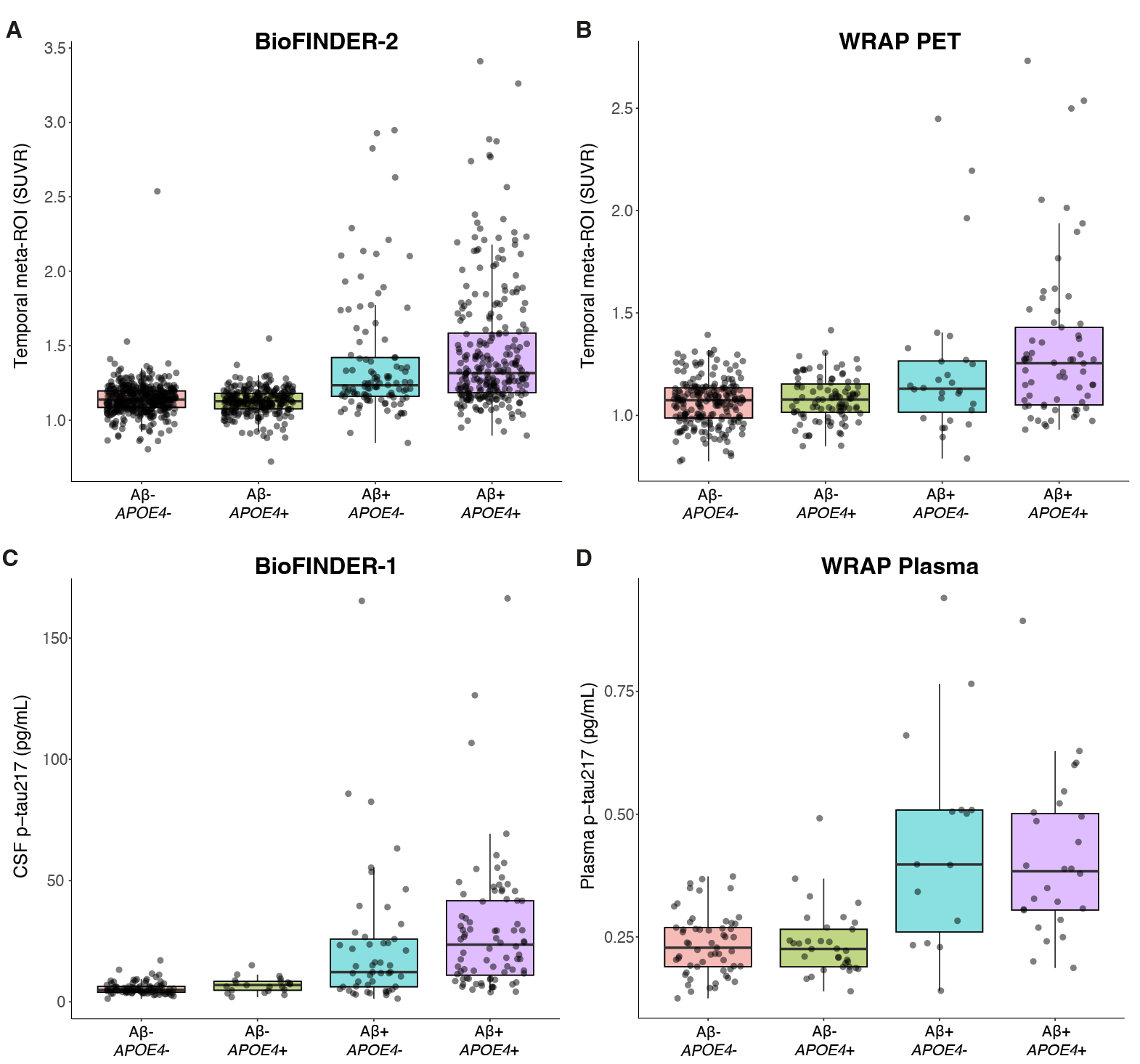

Boxplot for differences in baseline tau-PET SUVR between diagnostic groups defined by Aβ-PET status (-/+) combined with *APOE4* carriership (no ɛ4 allele, -; at least one ɛ4 allele, +). Tau-PET levels are shown in BioFINDER-2 (a) and WRAP PET (b). Differences based on *APOE4* carriership within Aβ+ and Aβ- groups were not significant (p=0.125-0.927). p-tau217 levels are shown in BioFINDER-1 (c) and WRAP (d). Differences based on *APOE4* carriership within Aβ+ and Aβ- groups were not significant (p=0.455-0.997). In all cohorts, the only differences were seen between the Aβ groups, with Aβ-positive participants had higher tau levels at baseline than Aβ-negative participants (p<0.001).

**Supplementary Figure 2. Mediation of Aβ on *APOE4* and temporal meta-ROI tau PET**

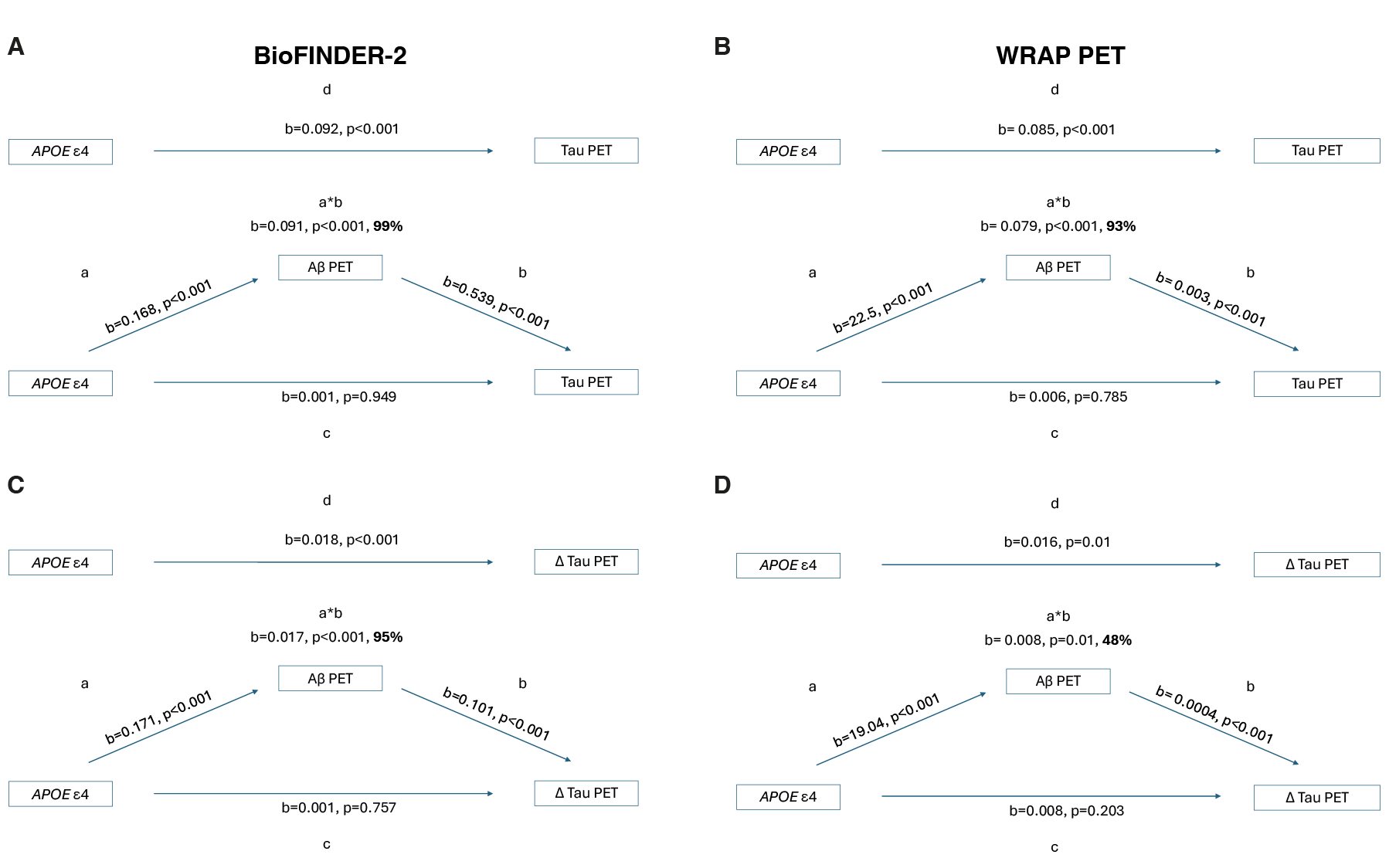
Mediation analysis for cross-sectional tau-PET in BioFINDER-2 (a) and WRAP (b) and longitudinal tau PET in BioFINDER-2 (c) and WRAP PET (d). Legend: a= effect of *APOE4* genotype on the mediator (Aβ-PET); b= effect of the mediator on tau, corrected by *APOE4* genotype; c= direct effect of *APOE4* on tau; d=total effect of *APOE4* on tau; a*b= mediation effect with % of the mediated effect on the total effect. *APOE4* had no significant direct effect on tau (a-d, p=0.203-0.949). *APOE4* effects on tau are fully mediated by Aβ.

**Supplementary Figure 3. Mediation of Aβ on *APOE4* and entorhinal tau PET**

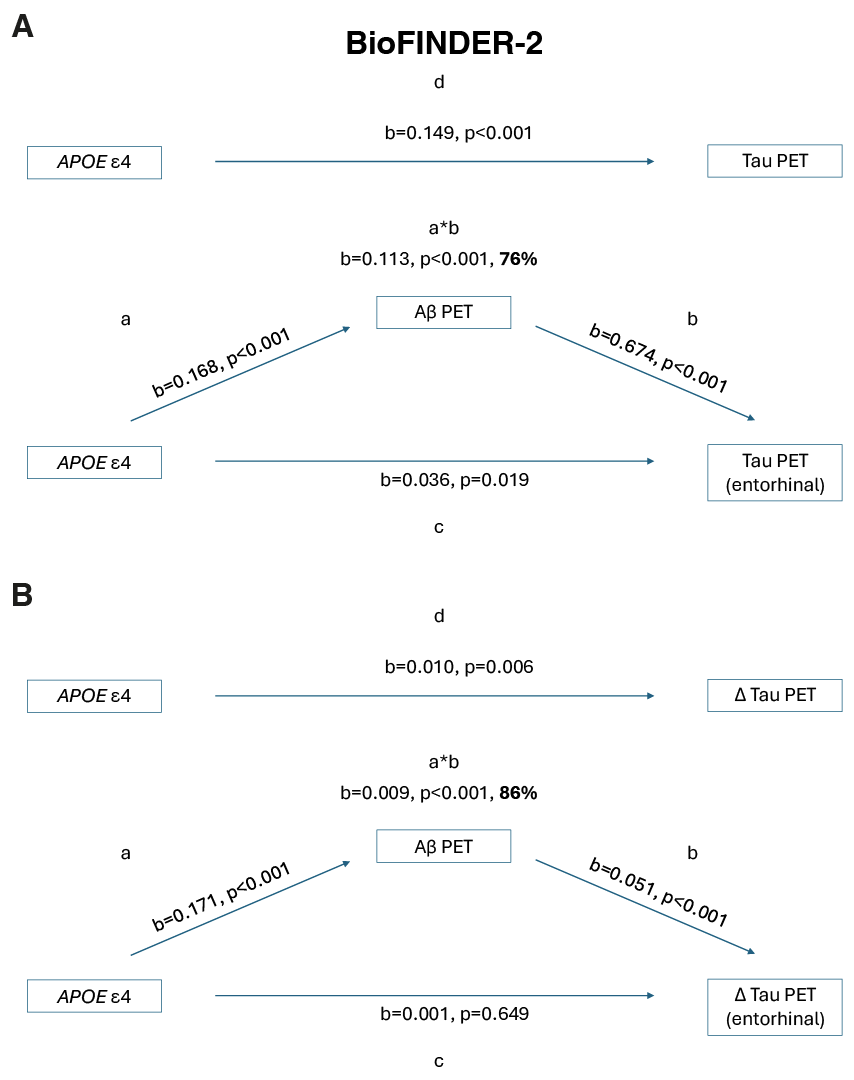

Mediation analysis for entorhinal tau-PET in BioFINDER-2 cross-sectionally (a) longitudinally (b). Legend: a= effect of *APOE4* genotype on the mediator (Aβ-PET); b= effect of the mediator on tau, corrected by *APOE4* genotype; c= direct effect of *APOE4* on tau; d=total effect of *APOE4* on tau; a*b= mediation effect with % of the mediated effect on the total effect. *APOE4* effects on tau over time are fully mediated by Aβ, but partially mediated by Aβ on baseline entorhinal tau.

**Supplementary Figure 4. Mediation of Aβ on *APOE4* and soluble p-tau217**

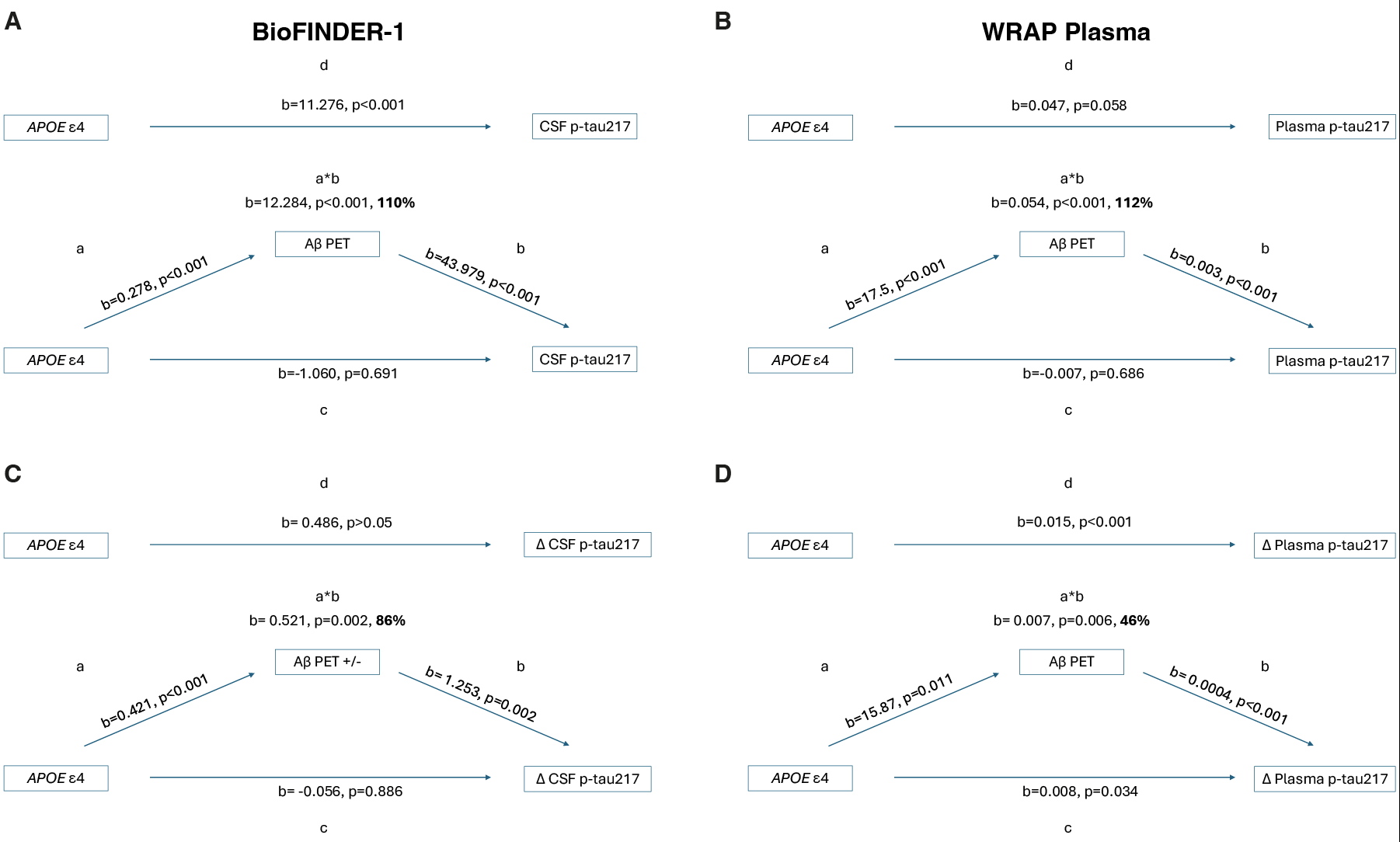

Mediation analysis for cross-sectional CSF p-tau217 in BioFINDER-1 (a) and plasma p-tau217 in WRAP (b) and longitudinal p-tau217 in BioFINDER-1 (c) and WRAP (d). Legend: a= effect of *APOE4* genotype on the mediator (Aβ-PET or Aβ-PET status +/-); b= effect of the mediator on tau, corrected by *APOE4* genotype; c= direct effect of *APOE4* on tau; d=total effect of *APOE4* on tau; a*b= mediation effect with % of the mediated effect on the total effect. *APOE4* has no significant direct effect on baseline/longitudinal CSF and baseline plasma tau (a-c, p=0.686-0.691), while it shows a significant effect on the rate of change of plasma tau (d, p=0.034). Continuous Aβ-PET SUVR in BioFINDER-1 showed no significant effect on longitudinal CSF p-tau217 (not shown), therefore we tested binary Aβ-PET status as mediator.

**Supplementary Figure 5. Interaction between *APOE4* based on the number of ε4 alleles and Aβ on tau-PET and p-tau217**

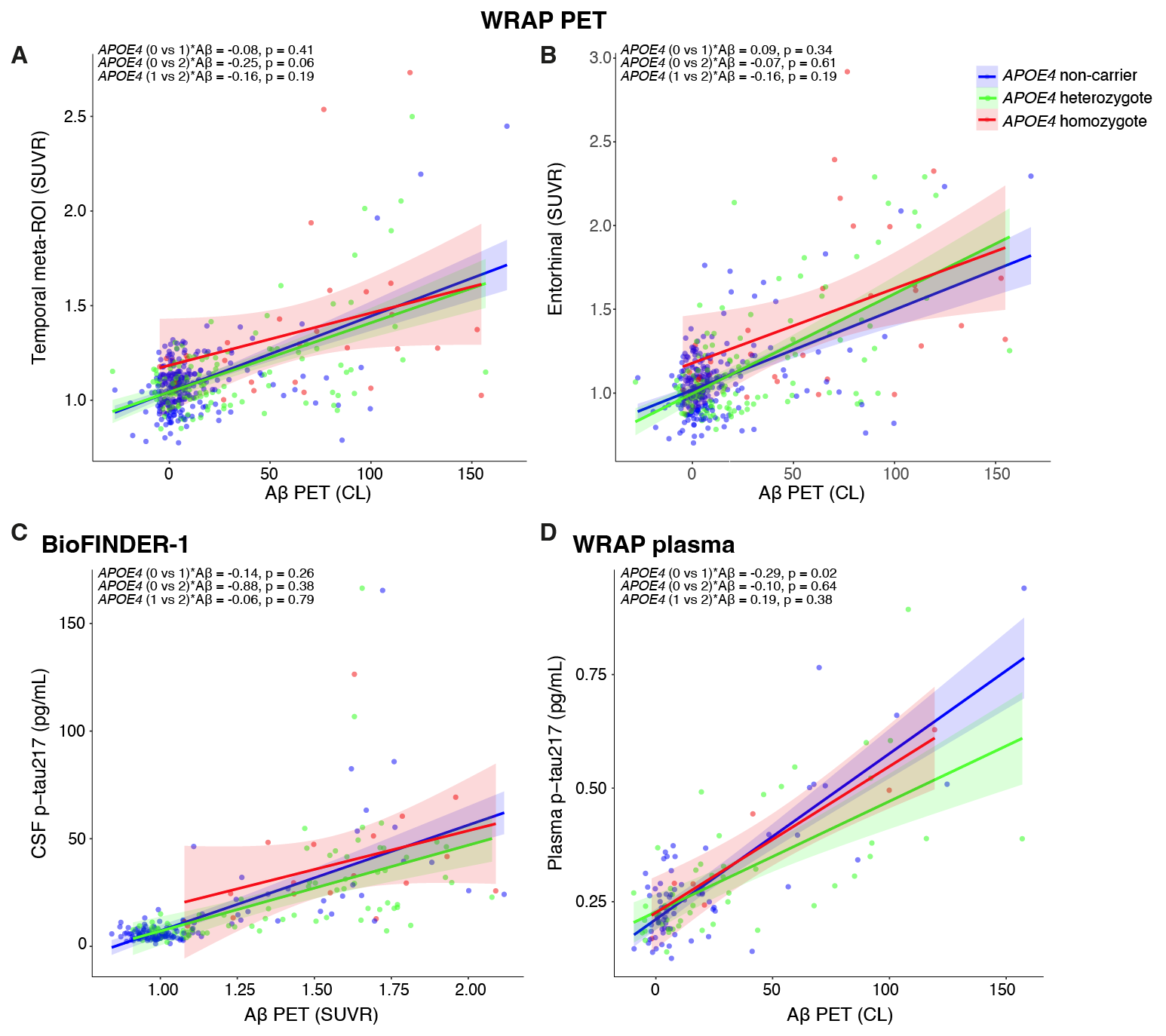

Interaction between *APOE4* groups and Aβ-PET on cross-sectional tau-PET in WRAP (a-b), CSF p-tau217 in BioFINDER-1 (c) and plasma p-tau217 in WRAP plasma (d). The *APOE4* groups are coded based on the number of alleles: 0 corresponds to non-carriers, 1 corresponds to *APOE4* heterozygotes; 2 corresponds to *APOE4* homozygotes. There were no significant interactions, but we note that APOE4 heterozygotes and homozygotes are small compared to the BioFINDER-2 cohort shown in Figure 3.
